# Spatial and temporal dynamics of SARS-CoV-2 in COVID-19 patients: A systematic review

**DOI:** 10.1101/2020.05.21.20108605

**Authors:** Anne Weiss, Mads Jellingsø, Morten Otto Alexander Sommer

## Abstract

**Background:** The spatial and temporal dynamics of SARS-CoV-2 have been mainly described in form of case series or retrospective studies. In this study, we aimed to provide a coherent overview from published studies of the duration of viral detection and viral load in COVID-19 patients, stratified by specimen type, clinical severity and age.

**Method:** We systematically searched PubMed/MEDLINE and Cochrane review database for studies published between 1. November 2019 and 23^rd^ of April 2020. We included studies that reported individual viral data over time measuring negative conversion by two consecutive negative tests, individual clinical severity and age. We excluded studies that reported viral data as patient fraction, reported only baseline data, included solely asymptomatic patients or were interventional studies. Extracted data included author, title, design, sample size, thresholds and genes of RT-PCR, patient age, COVID-19 severity, clinical characteristics, treatment, location of viral sampling, duration of viral detection, and viral load. We pooled the data of selected studies to determine effect estimates of duration of viral detection. Combined viral load was visualized over time.

**Findings:** Out of 7226 titles screened, 37 studies met the inclusion criteria and were included in the qualitative analysis and 22 studies in the quantitative analysis comprising 650 COVID-19 patients. The pooled estimate of the duration of positive detection of the virus was in mild adult patients 12.1 days (CI: 10.12, 14.05) after symptom onset in the upper respiratory tract (URT), 24.1 days (CI: 10.02, 38.19) in lower respiratory tract (LRT), and 15.5 days (CI: 8.04, 22.88) in faeces. Further, in mild adult patients, the maximum viral load was ~ 6.61 × 10^8^ viral copies/mL in the URT and ~ 2.69 × 10^8^ viral copies/mL in the LRT, within the first week of symptom onset. The maximum viral load in faeces was reported as ~ 3.55 × 10^7^ copies/mL on Day 9. In moderate-severe adult patients, the pooled estimate of mean duration of positive viral detection in the URT was 15.8 days (CI: 11.12, 20.56) after symptom onset, 23.2 days (CI: 21.49, 24.97) in the LRT, 20.8 days (CI: 16.40, 25.17) in faeces. The maximum viral load was 4.60 × 10^9^ copies/mL on Day 8 in the URT, 3.45 × 10^8^ copies/mL on Day 11 in the LRT, 2.76 × 10^6^ copies/mL on Day 18 in faeces and 1 × 10^4^ copies/mL on Day 3 in blood. In children with mild symptoms, the pooled estimate of the mean duration of positive SARS-CoV-2 viral detection was 11.1 days (CI: 7.14, 15.11) in the URT and 16.0 days (CI: 11.49, 20,47) in the faeces, without reporting quantitative viral data. Viral positivity was detected in the urine and eye in one patient.

**Interpretation:** Our analysis showed consistent viral detection from specimen from the URT, the LRT and faeces, irrespective of the clinical severity of COVID-19. Our analysis suggests that SARS-CoV-2 persists for a longer duration in the LRT compared to the URT, whereas the differences in the duration of viral detection between mild and moderate-severe patients is limited in the LRT, but an indication of longer duration of viral detection in feces and the URT for moderate-severe patients was shown. Further, viral load was demonstrated to peak in the URT within first weak of infection, whereas maximum viral load has been observed to occur later and within the second week of infection in the LRT.

**Funding:** The project has received funding support from Innovation Fund Denmark.

## Introduction

Coronavirus disease 2019 (COVID-19) has caused many deaths and severe suffering worldwide prompting a surge in research and drug development activities. These efforts are revealing clinical and molecular characteristics of SARS-CoV-2 and the infection it causes^1^, but its viral dynamics and spatial shedding patterns in humans remain poorly understood. A variety of case series and retrospectives studies, reporting viral dynamics of SARS-CoV-2 on a patient or patient population level, have been published and illuminate the spatial patterns of SARS-CoV-2’s shedding in different stages of the infection.^1^ However, these studies are heterogeneous with regards to patient populations and the specific viral load testing. To provide an overview of our current knowledge of the shedding patterns of SARS-CoV-2, we conducted a review of the temporal and spatial viral dynamics across clinical severities of COVID-19. Firstly, this knowledge can help improve mathematical models of SARS-CoV-2’s replication, as the numerical model can be trained with more real-world data. Secondly, this knowledge can inform the definition of relevant endpoints in clinical trials assessing pharmacological treatments aimed at reducing viral load in patients positive for SARS-CoV-2.

The objective of this paper is to provide a systematic review of the spatial and temporal viral dynamics of SARS-CoV-2 in COVID-19 patients, stratified after clinical severity, location of viral sampling and age, based on individual patient data. The data of the systematic review is then aggregated to report a weighted mean of duration of viral detection, per sampling location for adults and children with mild and moderate-severe symptoms.

## Methods

### Data sources

We searched the databases MEDLINE/PUBMED and Cochrane Review with the following search terms: “SARS-CoV-2 [MESH]” OR “COVID 19 [MESH]” alone, or in combination with “virology” OR “viral” OR “Epidemio*” AND “clinical”. Studies from November 2019 until 23^rd^ of April 2020 are included in this review.

### Eligibility criteria

The inclusion criteria with the corresponding rational is summarized in Table 1. Studies which demonstrated viral data as patient fractions (e.g. as Kaplan Meyer Plots), missed crucial information needed for detailed stratification (e.g. age, clinical severity, individual viral load/dynamic data), contained only asymptomatic patients or could be classified as interventional study were excluded.

**Table 1:**
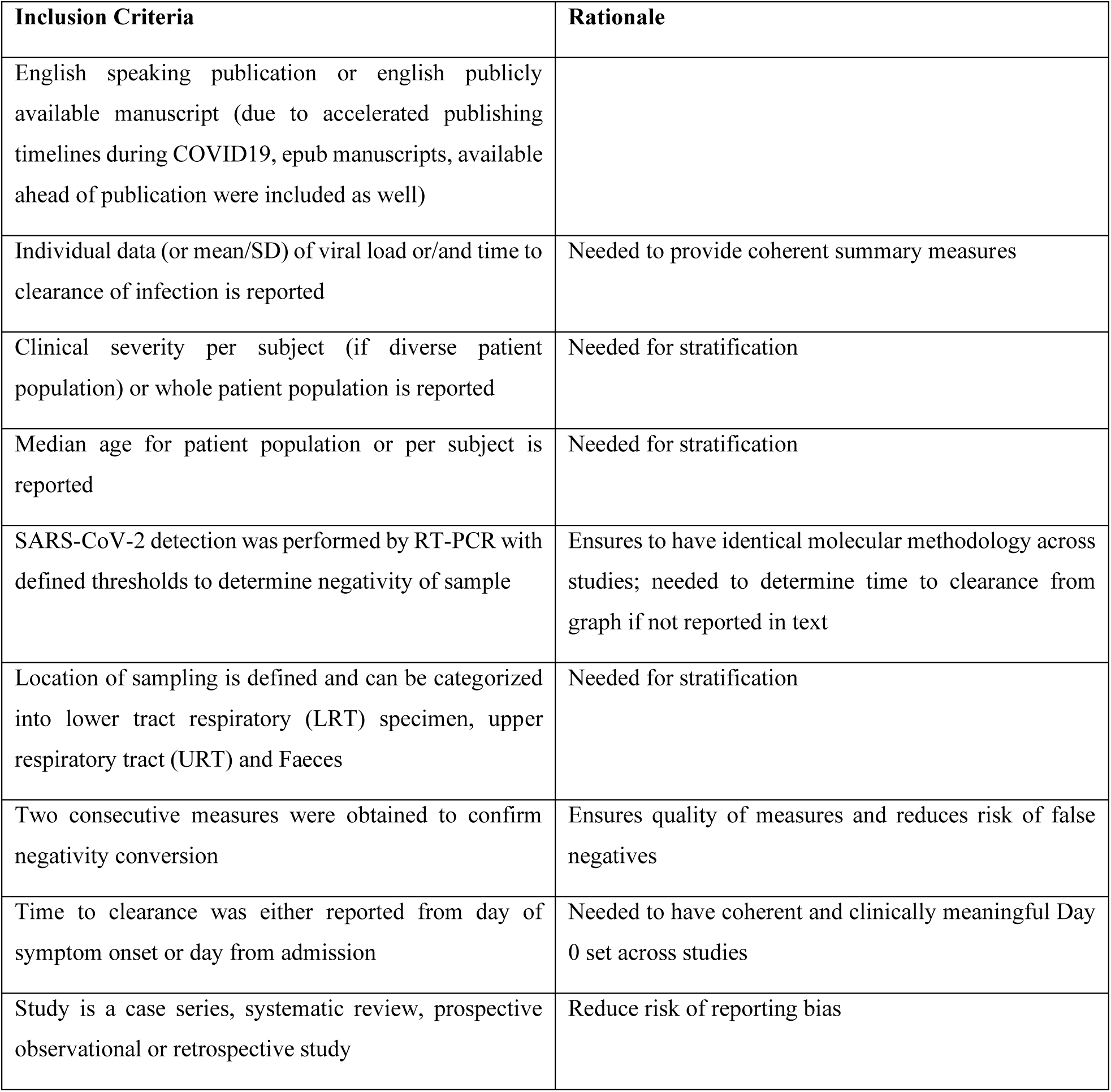
Inclusion criteria for eligibility of studies.

| Inclusion Criteria | Rationale |
| --- | --- |
| English speaking publication or english publicly available manuscript (due to accelerated publishing timelines during COVID19, epub manuscripts, available ahead of publication were included as well) |  |
| Individual data (or mean/SD) of viral load or/and time to clearance of infection is reported | Needed to provide coherent summary measures |
| Clinical severity per subject (if diverse patient population) or whole patient population is reported | Needed for stratification |
| Median age for patient population or per subject is reported | Needed for stratification |
| SARS-CoV-2 detection was performed by RT-PCR with defined thresholds to determine negativity of sample | Ensures to have identical molecular methodology across studies; needed to determine time to clearance from graph if not reported in text |
| Location of sampling is defined and can be categorized into lower tract respiratory (LRT) specimen, upper respiratory tract (URT) and Faeces | Needed for stratification |
| Two consecutive measures were obtained to confirm negativity conversion | Ensures quality of measures and reduces risk of false negatives |
| Time to clearance was either reported from day of symptom onset or day from admission | Needed to have coherent and clinically meaningful Day 0 set across studies |
| Study is a case series, systematic review, prospective observational or retrospective study | Reduce risk of reporting bias |

### Data extraction

Data were extracted from the full text/supplemental material or if only available in graphs digitalized using GetData Graph Digitizer © version 2.26.0.20. We used a pre-defined extraction protocol to capture identical information across studies (incl. author, title, country, design, sample size, thresholds and genes of RT-PCR, frequency of sampling, patient age (median), COVID-19 severity and classification reference, major clinical characteristics, treatment, location of viral sampling, duration of viral detection, viral load).

### Data processing and summary measures

To determine the duration of viral detection from symptom onset, we counted the days from initiation of symptoms (Day 0) to the last positive test (final day was set as Day-1 of first negative test of two consecutive tests for daily sampling). However, in some studies the sampling frequency was not daily, meaning that the last positive test does not equal Day-1 of first negative test. Therefore, we also reported the duration of viral detection until the last positive swab, to not introduce any predictions into the summary measures. In the case that the day of the positive test was not reported for a patient, we excluded this patient in the overall summary measure, as the duration of viral detection becomes predictive. Also, in case a positive test was not reported, we could not be sure that this patient was tested at all positive for the virus at the specific sampling location. Furthermore, for patients that did not clear the virus within the time frame of sampling, we used the day of the last positive test as final day to not introduce bias into the data by excluding these and leaving out patients that needed longer to clear the virus than the average. Finally, we also recorded the duration of viral detection in case recurrence of positivity was observed, meaning a positive test was detected after two negative ones have been obtained. To measure the duration in regard to the “recurrence” endpoint, the final day was also set as Day-1 of the first negative test of two consecutive negative tests.

In case clinical severity was not categorized into mild, moderate or severe by the authors of the publication, we conducted this categorization with the symptoms reported, based on definitions described by Wu *et al.*. ^2^Asymptomatic patients were excluded.

Duration of viral detection is demonstrated as mean (days after symptom or admission onset) with its corresponding standard deviation and viral load was visualized as viral copies/mL [log10] over time (days after symptom onset) stratified after clinical severity, mild or moderate/severe, and location of swab (upper respiratory tract (URT) specimen, lower respiratory tract (LRT) specimen or faeces). Oro-/nasopharyngeal, throat and nasal swabs were combined to “URT”, sputum and tracheal aspirate to “LRT”, anal swabs and stool to “faeces”, and plasma and serum to “Blood”. Data which did not use copies/mL as unit for quantifying viral load were excluded. R Studio ©, Version 1.2.1335 was used for visualization. Raw data can be found in Supplementary Table S1.

### Statistical analysis

Statistical analysis comparing the average load per sampling location across each study was performed using unpaired t test, not assuming consistent SD and using a two stage setup false discovery rate approach of Benjamini, Krieger and Yekutieli, setting the desired FDR to 1%. ^3^ Statistical analysis was conducted in GraphPad Prism 8.1.2.

### Aggregation of data

#### Study Selection

We included studies measuring duration positive detection of SARS-CoV-2 in days after symptom onset, but not after hospitalization ensuring that an identical Day 0 is set across studies. Patients equal to 18 years or older were classified as adults (if only median was recorded, the median age was considered for categorization). Single case series were excluded. In total, 22 Studies were included in the aggregation of data. ^4–25^

#### Data processing

Oro-/nasopharyngeal throat and nasal swabs were combined to “URT”, sputum and tracheal aspirate to “LRT”, anal swabs and stool to “faeces”, and plasma and serum to “Blood”. Mild disease severity was kept as mild and moderate and severe disease severity were merged to moderate-severe in aggregation analysis.

#### Statistical analysis

Aggregation of study data of duration of viral detection was conducted in ReviewManager 5.3. Standard error was computed according to STDEV/(SQRT(N)). We used generic inverse variance as a data type (mean and SE), and random effect as analysis model. Effect measures were shown as mean with 95% confidence interval (CI). Heterogeneity was measured by I^2^. Forest plots are provided for selected data groupings.

## Results

### Study Selection

The overall study selection process is displayed in the flow-diagram in Figure 1. Briefly, we screened a total of 7226 titles and selected 553 for abstract screening. Out of the 553 abstracts, we selected 143 studies to be scanned as full text to investigate whether viral data is generally reported. Out of these studies, 80 studies were chosen for in-depth reading to assess eligibility. Finally, 37 studies were included in the systematic review. Reasons for exclusion were: individual patient data was missing and/or only median of duration of viral detection was demonstrated, clinical severity or age was not reported or only baseline viral data were shown.

**Figure 1:**
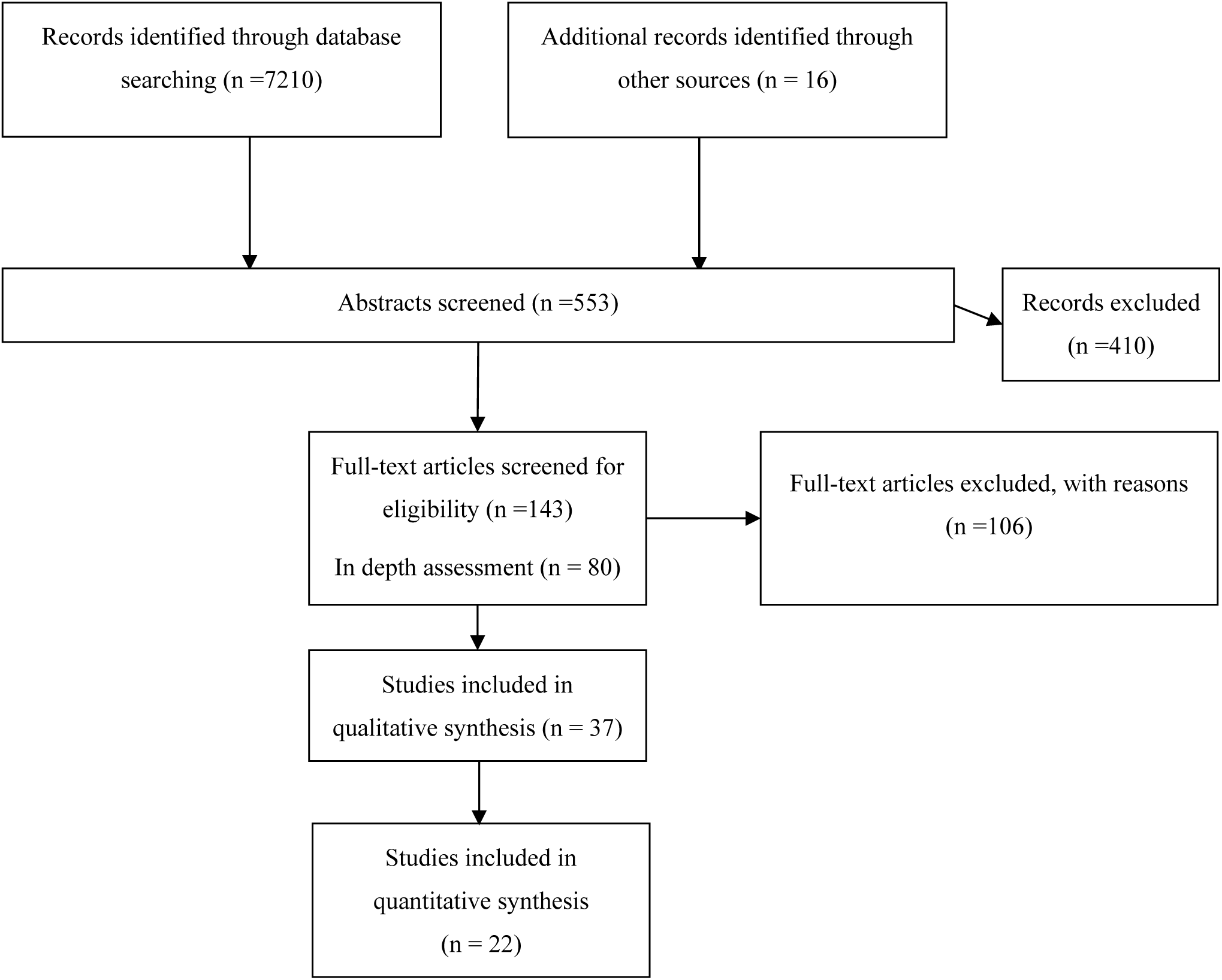
Study Selection.

### Overview of studies describing duration of viral detection in COVID-19 patients

The data summary of the selected studies, describing duration of viral detection as days after symptom onset (as mean and SD) is displayed in Table 2 and days after hospitalization in Table 3. More details of each study, including clinical characteristics can be found in Table S2 in the supplementary material.

**Table 2:**
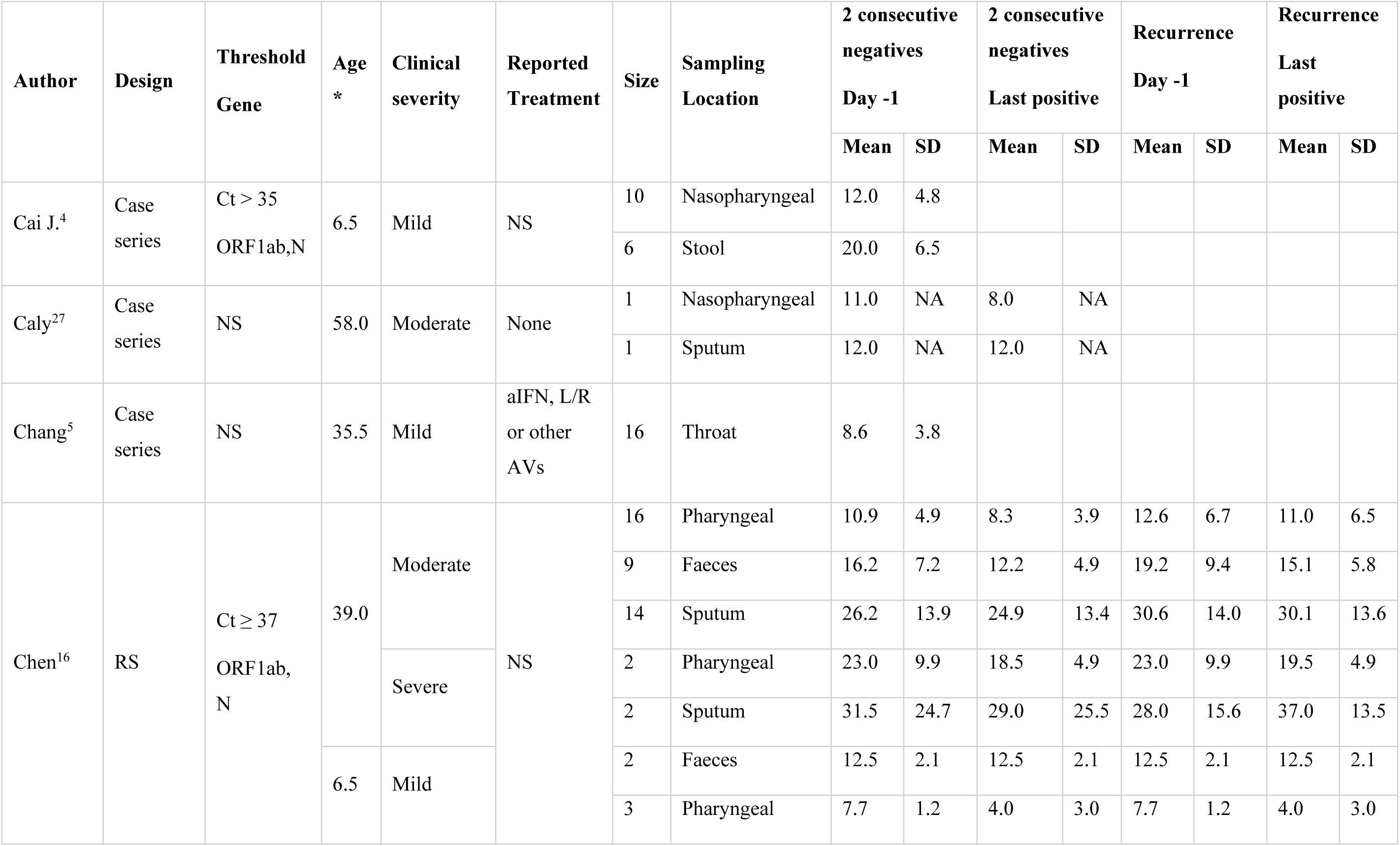
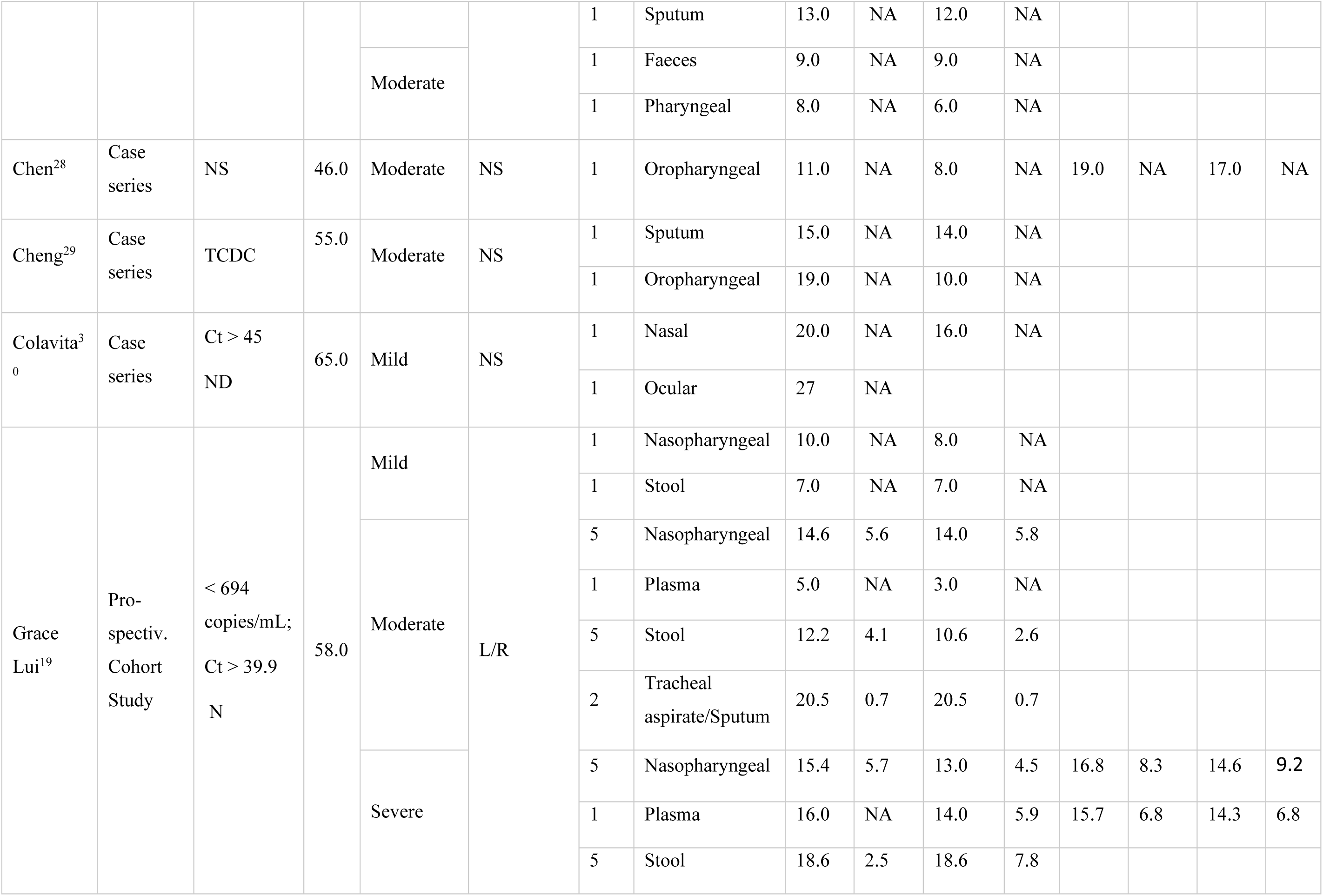
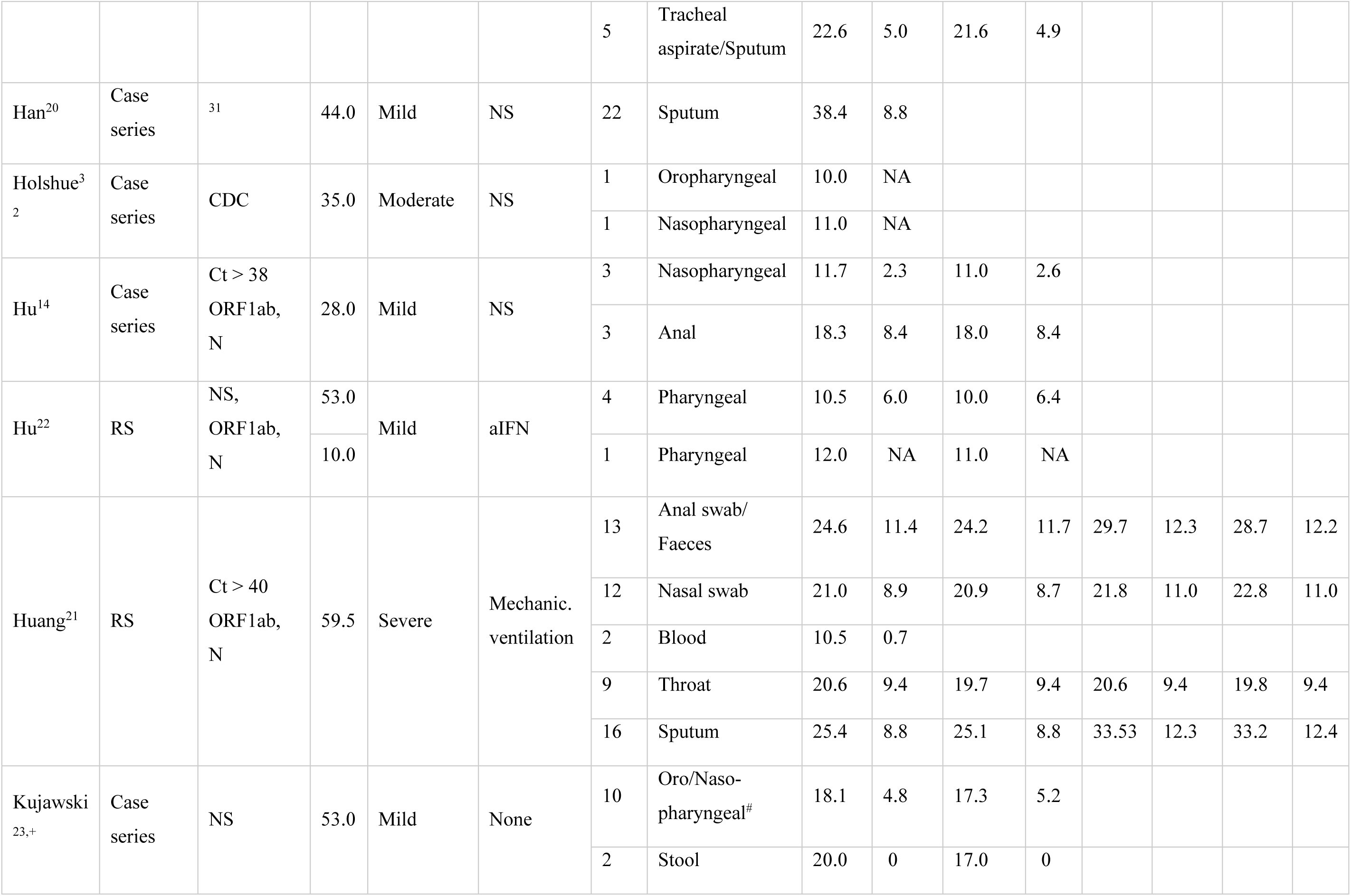
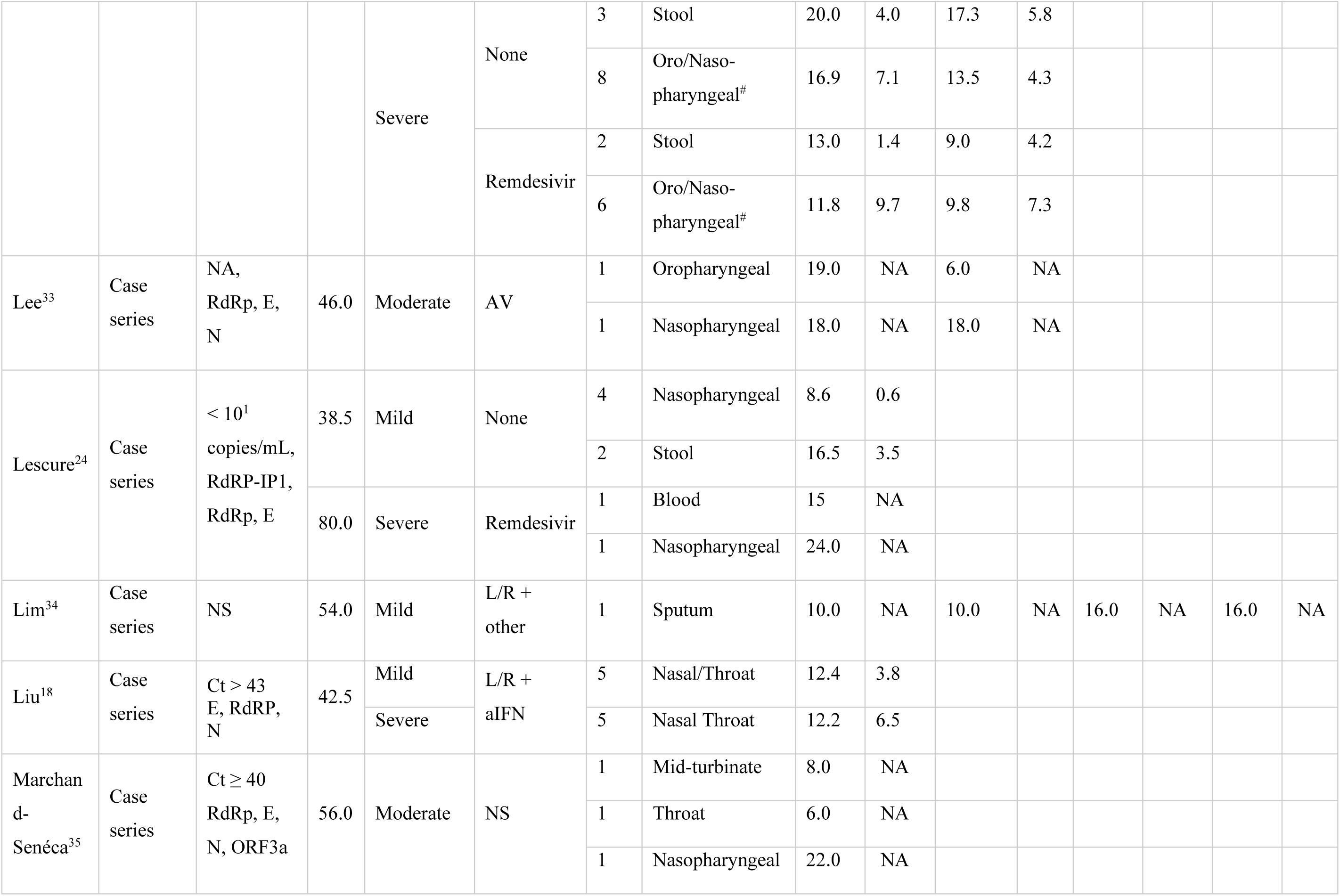
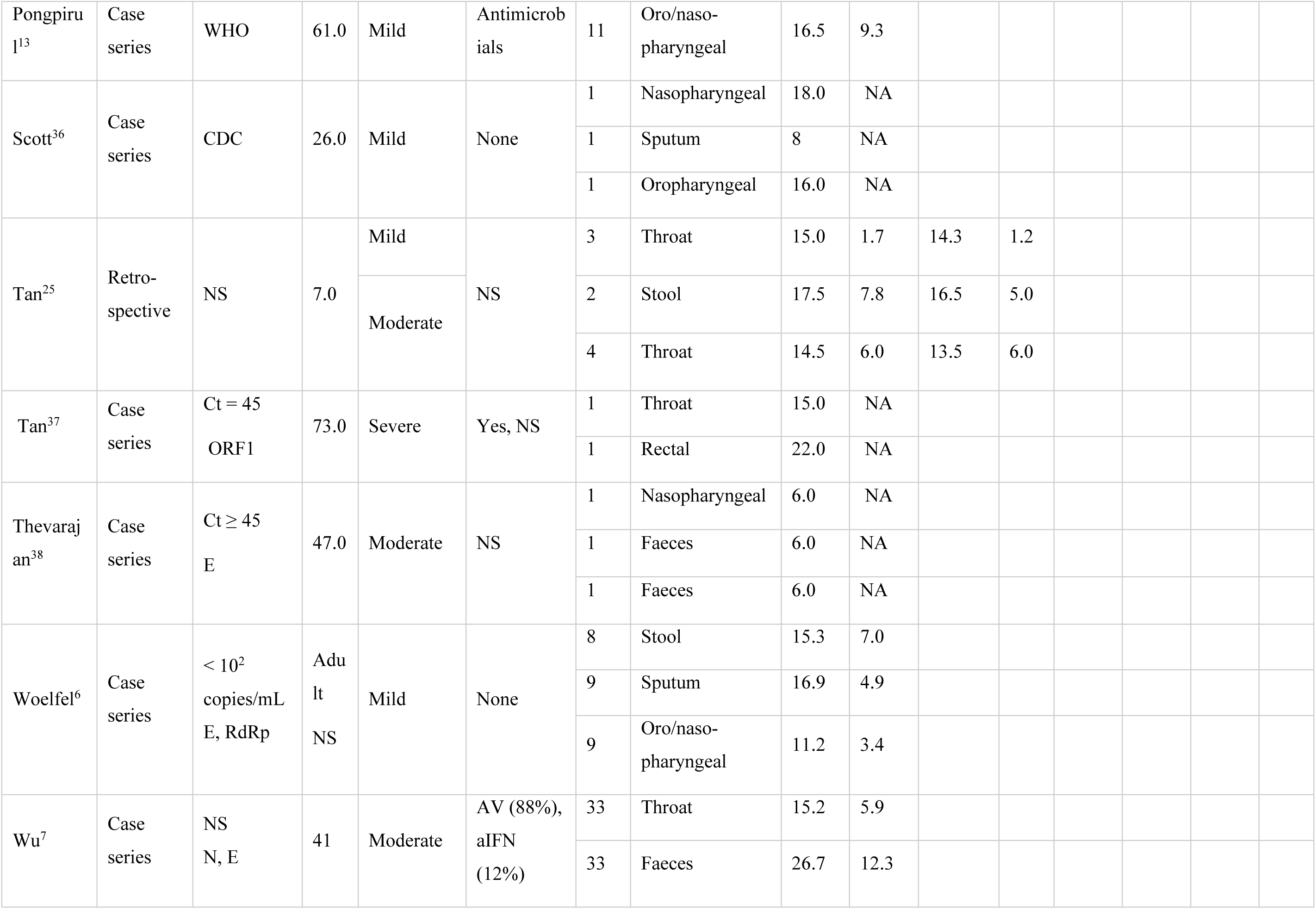
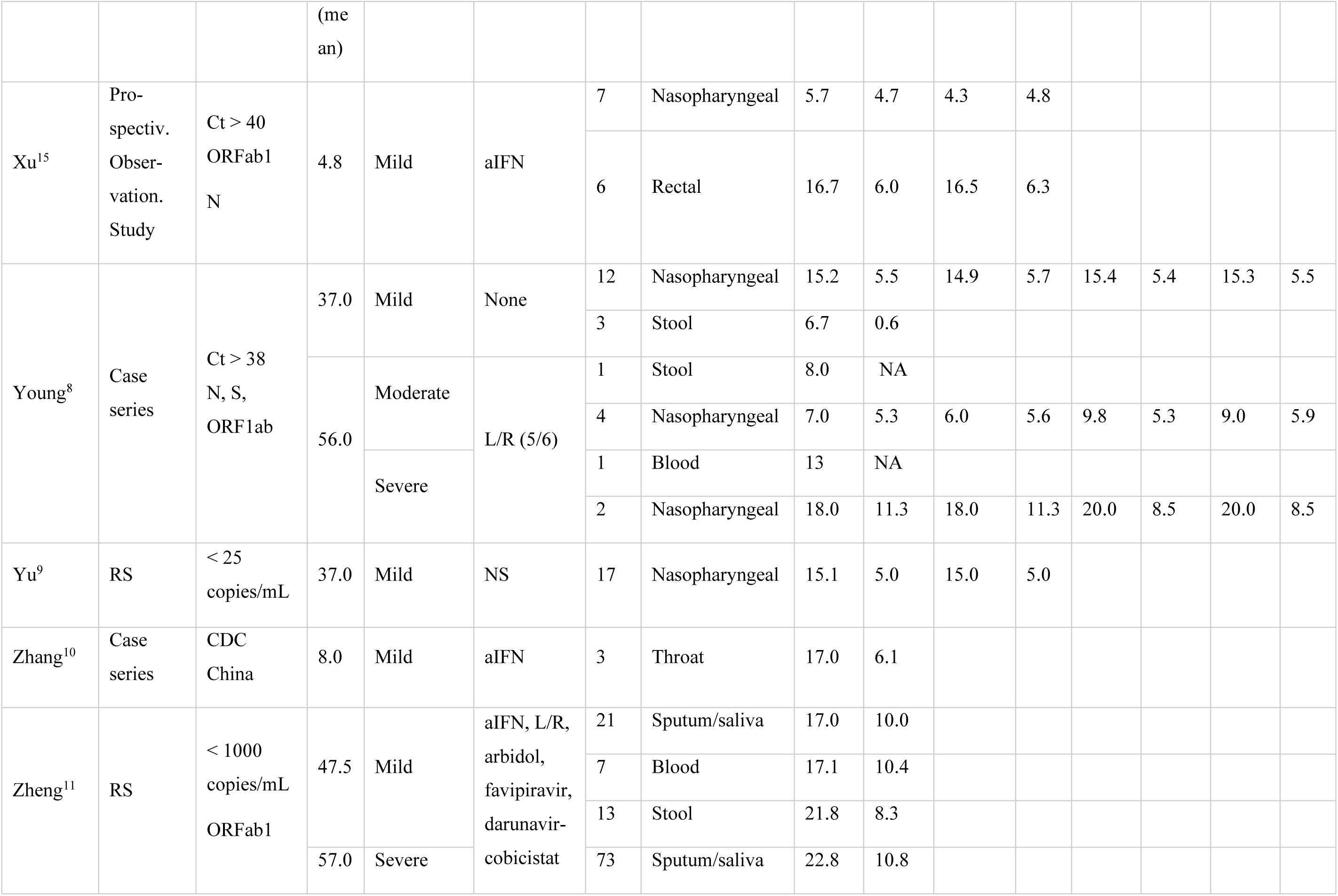
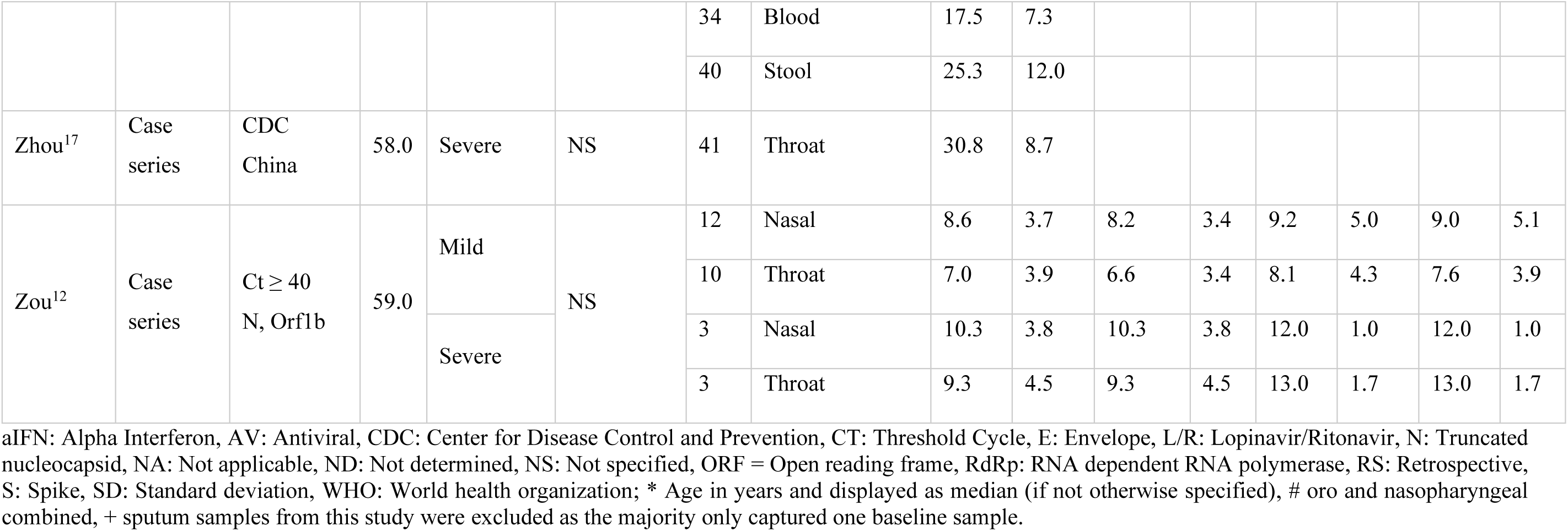
Summary of studies describing duration of viral detection as days after symptom onset. Blank cells apply if sampling was daily (value of Day-1 equals value of last positive) or no recurrence occurred. Samples that were not tested positive are excluded in this overview.

**Table 3:**
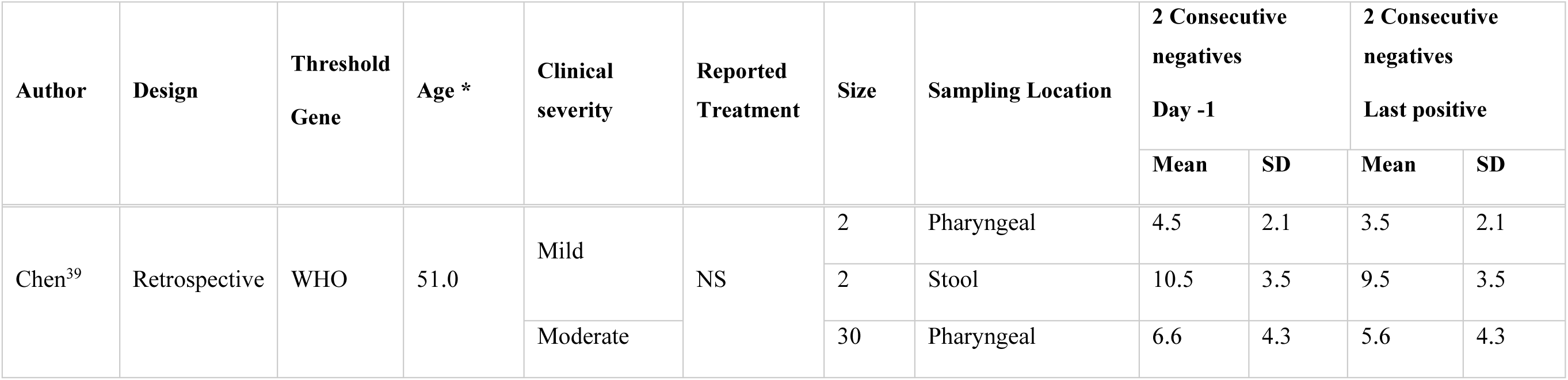
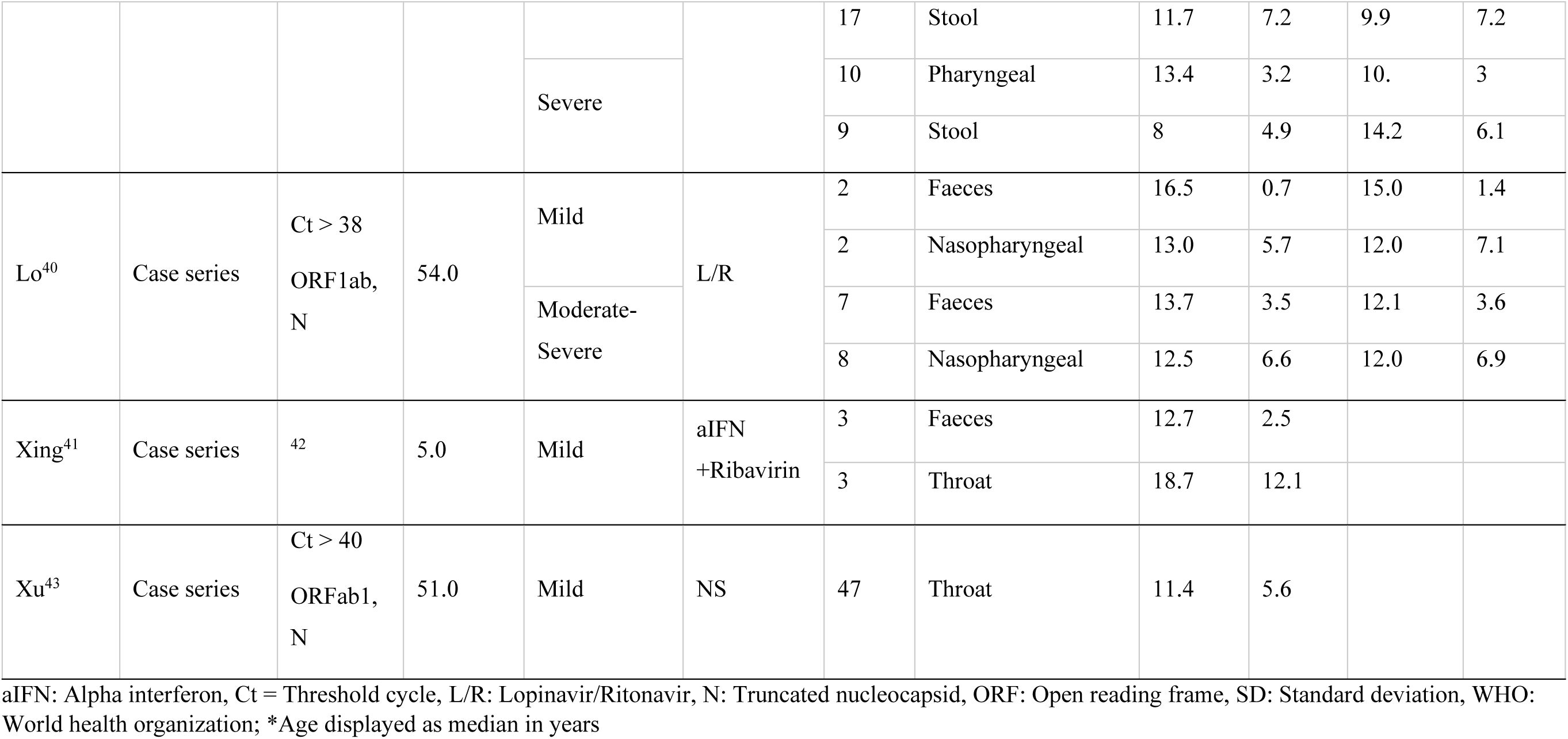
Summary of studies describing duration of viral detection as days after hospitalization. No recurrence was reported for these studies. Blank columns apply if sampling was daily (value of Day-1 equals last positive) or no recurrence occurred.

The SARS-CoV-2 virus is detected in both in the lower and upper respiratory tract, and faeces, irrespective of the severity of the disease.

The average duration of positive SARS-CoV-2 viral detection (Day-1) from URT specimens ranged from 7.9 days to 20 days after symptom onset in mild adult patients, and from 6 to 30.8 days in moderate-severe patients. The average duration of positive SARS-CoV-2 viral detection from LRT specimens ranged from 8 days to 38.4 days after symptom onset in mild adult patients, and from 6 days to 26.9 days in adults with moderate-severe symptoms. In children with mild COVID-19 symptoms, average duration of positive SARS-CoV-2 viral detection ranged from 5.7 days to 17 days after symptom onset in the URT and was reported as 13 days in the LRT in one study^16^. Duration of positive SARS-CoV-2 viral detection in children with moderate symptoms was observed in two studies reporting viral detection for 8 days and 14.5 days after symptom onset in the URT. ^16, 25^ Furthermore, several studies have reported positive SARS-CoV-2 viral detection in the faeces and blood in adults, and in faeces only in children. One adult patient was reported to have viral positivity in ocular fluids^30^, whereas other studies did not find evidence of positive SARS-CoV-2 viral detection in the eye. ^9,23,24^ Only one adult patient was reported to have a positive viral sample in the urine. ^21^

Recurrence of viral positivity after two consecutive negative tests were obtained, was observed in 7 out of 37 studies. Recurrence of positivity occurred in adult patients and URT specimen with mild symptoms (1 patient in LRT), and in mild-severe adult patients and URT, LRT and faecal specimen.

Generally, the average duration of viral detection only differed minimally when setting the final day as Day-1 of first negative test (results described in text above) or as day of last positive test in the above patient populations.

### Aggregation of study data

We aggregated the data of 22 selected studies to report a weighted mean of the duration of positive SARS-CoV-2 viral detection. We report the weighted mean for both outcome measures, negative conversion defined by obtaining two consecutive negative samples and negative conversion after recurrence (positive sample was detected after two consecutive negatives were obtained). As sampling frequency varied and was not daily for all the studies, we also report the results obtained when setting different endpoint measures: 1) final day of positive detection of virus is day-1 of first negative of two consecutive measurement and 2) final day of positive detection of virus is the day on which last positive test was obtained, in case Day-1 and last positive day were not identical, to avoid introducing predictions into the outcome measures. Notably, there was a portion of patients that did not reach viral negativity within the time of sampling for which we reported the day of the last positive test as final day. Considering studies that were included in the aggregation analysis, the share of mild patients (children and adults) that did not reach negativity were 27/139 for URT specimen, 6/52 for LRT specimen and 26/45 for faecal specimen. Regarding moderate-severe patients (children and adults), these were 14/154 for URT specimen, 6/112 for LRT specimen, 0/38 for blood specimen and 10/110 for faecal specimen. For the overall patient population, the patients that did not clear within the sampling time accounted for 14% of the whole dataset.

We aggregated the data of selected studies to report a weighted mean of duration of positive SARS-CoV-2 viral detection in both URT and LRT for mild and moderate-severe patients. We chose to display the results of the aggregated data for which the final day was set as Day-1 of first negative in the main text of this manuscript, as this is the most conservative approach (Figure 2 and Figure 3). Also, the result of this measure differed only marginally to the result defining the final day as last positive (maximum 1 Day). The pooled estimates for all data groupings, including recurrence, can be found in the Supplementary Table S3.

**Figure 2:**
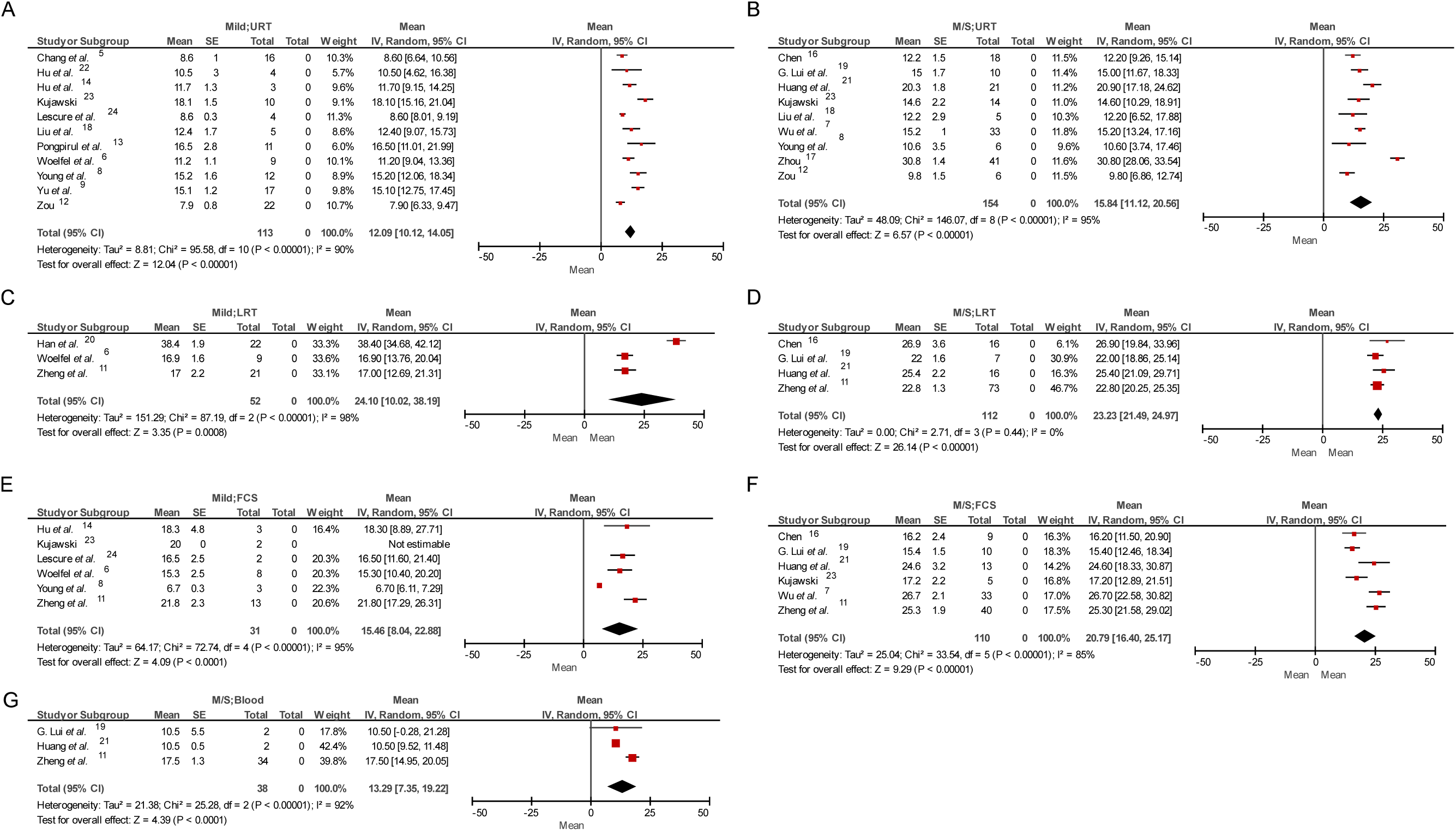
Forest plots of selected aggregations reporting weighted means of duration of viral detection as days after symptom onset in adults. Pooled estimate for (A) all mild patients (non-treated and treated) and URT specimens, (B) all moderate-severe patients and URT specimen, (C) all mild patients and LRT specimen, (D) all moderate-severe and LRT specimen, (E) all mild patients and faecal specimen, (F) moderate-severe and faecal specimen and (G) moderate-severe patients and blood specimen. The forest plots include data that describe duration of viral detection defining the final day as day-1 of first negative day of two consecutive negatives. CI: Confidence Interval; D-1: Day-1 of first negative test of two consecutive; FCS: Faeces, LRT: Lower respiratory tract, SE: Standard error, URT: Upper respiratory tract.

**Figure 3:**
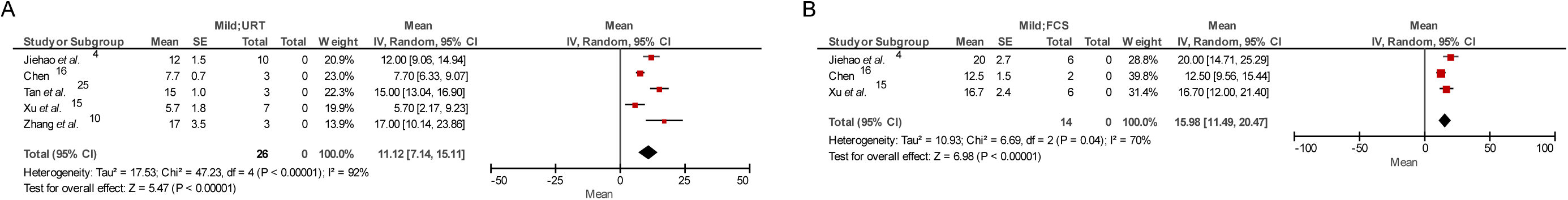
Forest plots of selected aggregations reporting weighted means of duration of viral detection as days after symptom onset in children. Pooled estimate for (A) all mild patients (non-treated and treated) and URT specimens, (B) all mild patients and faecal specimen. The forest plots include data that describe duration of viral detection defining the final day as day-1 of first negative day of two consecutive negatives. CI: Confidence Interval; D-1: Day-1 of first negative test of two consecutive; FCS: Faeces, SE: Standard error, URT: Upper respiratory tract.

In total we included data from 650 patients in this analysis. Notably, a high heterogeneity (I^2^ ~ 80-90%) was observed in the majority of the subgroup aggregation analysis indicating that these data should be interpreted cautiously and only considered as trends.

In mild adult patients, the pooled estimate of the mean duration of positive SARS-CoV-2 viral detection after symptom onset in the URT was 12.1 days (CI: 10.12, 14.05), 24.1 days (CI: 10.02, 38.19) in LRT, and 15.5 days (CI: 8.04, 22.88) in faeces (Figure 2A, 2C and 2E). There was only one study describing duration of viral detection in the blood (17.1 days) in mild adult patients^11^.

In moderate-severe adult patients, the pooled estimate of mean duration of positive SARS-CoV-2 viral detection after symptom onset in the URT was 15.8 days (CI: 11.12, 20.56) days, 23.2 days (CI: 21.49, 24.97) in the LRT, 20.8 days (CI: 16.40, 25.17) in faeces, and 13.3 days (CI: 7.35, 19.22) in blood (Figure 2B, 2D, 2F and 2G).

In children with mild symptoms, the pooled estimate of the mean duration of positive SARS-CoV-2 viral detection after symptom was 11.1 days (CI: 7.14, 15.11) in URT and 16.0 days (CI: 11.49, 20,47) in the faeces, (Figure 3). No viral detection was reported in the blood for children.

Generally, the mean duration of positive SARS-CoV-2 viral detection accounting for recurrence of positivity did not differ substantially in mild adults or children (if different, less than 1 Day difference). But, in moderate-severe patients the pooled estimate of the mean duration of positive SARS-CoV-2 viral detection was on average approximately 3.4 days longer for LRT specimen and 1.2 days for faeces, accounting for recurrence of viral positivity.

### Spatial and temporal dynamics of viral load

To investigate whether the viral load differs between disease severity and sampling location, we combined data from four published studies and analyzed the viral load per patient as a function of time, stratified after clinical severity (mild and moderate-severe) and location of sampling (URT, LRT and faeces) (Figure 4). No quantitative viral data was available for children and only one study reported limited quantitative data of blood specimen (four datapoints). ^19^

**Figure 4:**
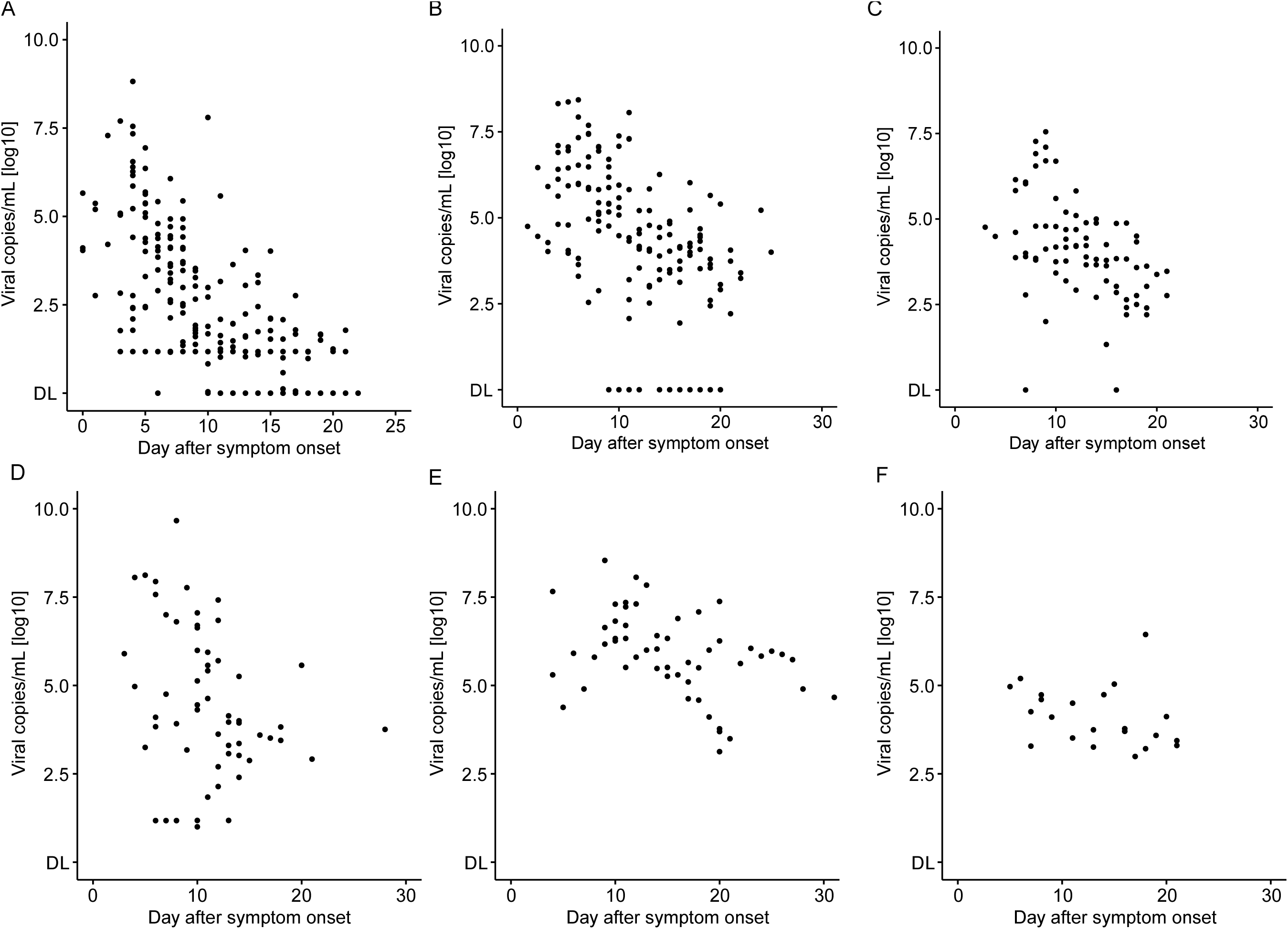
Spatial and temporal viral load in mild and moderate-severe adult COVID-19 patients. Temporal dynamic of viral detection in (A) URT specimen in mild patients, (B) LRT specimen in mild patients, (C) fecal specimen in mild patients, (D) URT specimen in moderate-severe patients, (E) LRT specimen in moderate-severe patients, (F) fecal specimen in moderate-severe patients. Four Studies were included^6,11,12,19^. DL: limit of detection.

**Figure 5:**
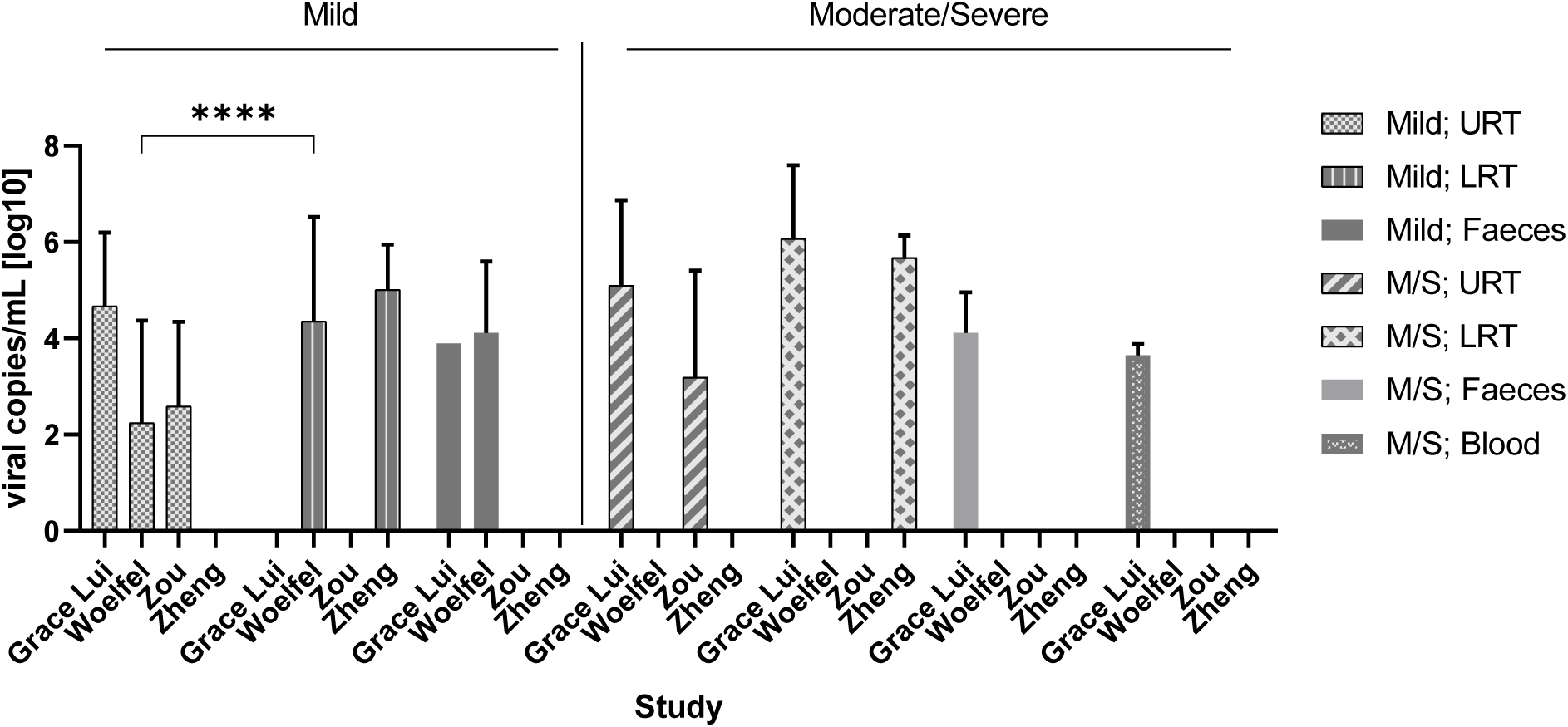
Average viral load per location and clinical severity. Bar graph displaying mean with SD. **** q < 0.001, unpaired t-test using false discovery approach of Benjamini, Krieger and Yekutieli; M/S: Moderate-severe, URT: upper respiratory tract, LRT: lower respiratory tract.

In mild patients, viral load was highest during the first week in the URT having a maximum viral load of ~ 6.61 × 10^8^ viral copies/mL on Day 4, whereas the maximum viral load reported for LRT was ~ 2.69 × 10^8^ copies/mL on Day 6 (Table 4). The maximum viral load in faeces was reported as ~ 3.55 × 10^7^ copies/mL on Day 9 in mild patients. The average viral load was found to be significantly higher in LRT relative to URT in the Woelfel *et al*. ^6^ study (t test, q < 0.001) (Figure 4). However, as this was the only study which allowed this comparison, additional studies of quantitative viral dynamics are needed to assess the viral load of the URT compared to LRT.

**Table 4:**
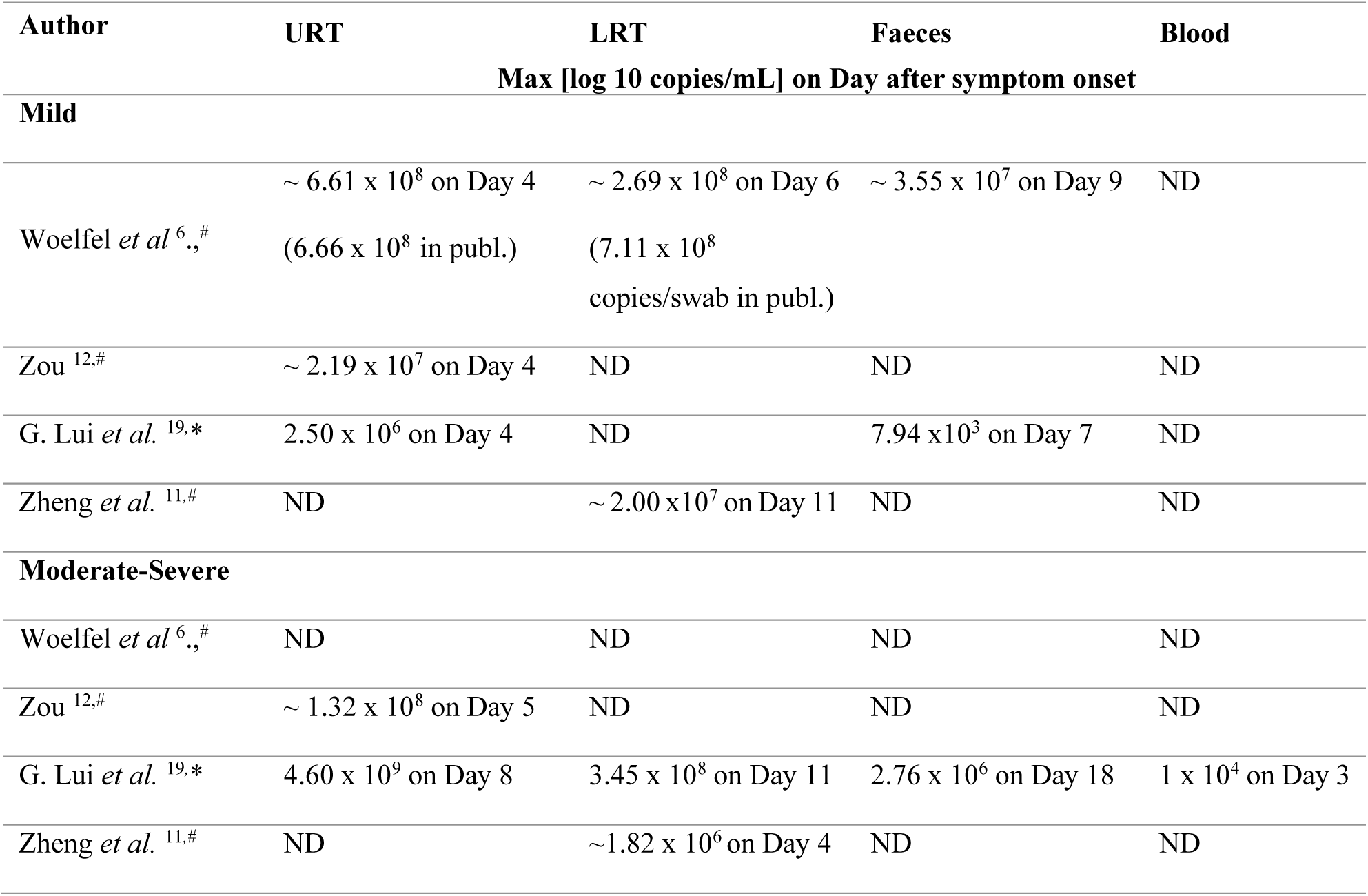
Summary of maximum viral load per location and COVID19 clinical severity. LRT: Lower respiratory tract, ND: Not determined, URT: Upper respiratory tract; * all subjects in this study received Lopinavir/Ritonavir; LRT: Lower respiratory tract, ND: Not determined, publ.: Publication; URT: Upper respiratory tract; # estimated data as digitalized from graph

In moderate-severe patients, the maximum viral load was reported as 4.60 × 10^9^ copies/mL on Day 8 in URT and 3.45 × 10^8^ copies/mL on Day 11 in LRT. Maximum viral load in faeces was shown to be 2.76 × 10^6^ copies/mL on Day 18 and 1 × 10^4^ copies/mL on Day 3 in blood specimen. No significant difference was found in the average viral load comparing URT and LRT samples in this disease population.

## Discussion

In this study we conduct a systematic review of the spatial and temporal viral dynamics of SARS-CoV-2 stratified after age and clinical severity. We report the per study data of duration of viral detection and viral load over time, but also show aggregated data of these, providing a better basis for interpretation. While viral trophism likely extends beyond the specimens described in this study^26^, the lack of data on viral load from other locations has limited our analysis to faeces, blood, URT and LRT.

Additionally, this study has several limitations which need to be considered. First, different genes and thresholds were used to assess negative conversion of SARS-CoV-2 hampering the direct comparison of studies. Second, the classification of clinical symptoms into the severity categories, mild, moderate, severe was based on different guidelines across the studies and we were not able to unify and verify all (mostly due to missing individual symptom data). Third, studies that only reported fractions or median of duration of viral shedding were excluded, which might introduce a selection bias into this review. Fourth, the viral load was measured via RT-PCR, which cannot differentiate dead virus particle, and hence data presented here might not necessarily reflect active viral replication. However, this technique is currently used worldwide to measure the quantities of SARS-CoV-2. Fifth, a portion of the patients included in this review (14 %) did not clear the virus in the time frame of sampling, hence the here presented data might be an underestimation of the duration of virus detection. Finally, our aggregation analysis exhibited a high heterogeneity (I^2^ ~ 80-90%), which highlights that these data should be interpreted cautiously and only considered as trends.

Regardless of these limitations, some trends can be extracted from this analysis. Firstly, we consistently find that SARS-CoV-2 is detected in LRT, URT and fecal specimens, irrespective of clinical severity of disease. Second, our data indicates that the duration of detection of SARS-CoV-2 is longer in LRT than the URT. Third, there seems to be little difference in the duration of detection of SARS-CoV-2 in mild patients as opposed to moderate-severe patients in the LRT, but an indication of longer duration of viral detection in feces and the URT for moderate-severe patients was shown. Fourth, viral load in URT peaks within the first week of infection, whereas viral load in LRT and faeces within the second week of infection.

In conclusion, we believe that this systematic review can support refinement of mathematical modelling as well as aid in the definition of appropriate endpoints for clinical trials tested therapeutic intervention aimed at reducing viral load of SARS-CoV-2. Furthermore, it highlights the importance of testing for viral presence in the LRT, which may clear more slowly compared to the URT.

## Data Availability

All data used for this manuscript were publicly available or provided by the author upon request.

## Contributors

A.W. and M.O.A.S wrote the manuscript and conceived the study. M. J. and M.O.A.S. provided input to the analysis and interpretation of study. A.W. analyzed the data. All authors reviewed the manuscript.

